# Digital gait outcomes for ARSACS: discriminative, convergent and ecological validity in a multi-center study (PROSPAX)

**DOI:** 10.1101/2024.01.04.24300722

**Authors:** Lukas Beichert, Winfried Ilg, Christoph Kessler, Andreas Traschütz, Selina Reich, Filippo M. Santorelli, Ayşe Nazli Başak, Cynthia Gagnon, PROSPAX consortium, Rebecca Schüle, Matthis Synofzik

**Affiliations:** Division Translational Genomics of Neurodegenerative Diseases, Hertie-Institute for Clinical Brain Research and Center for Neurology, University of Tübingen, Tübingen, Germany; German Center for Neurodegenerative Diseases (DZNE), University of Tübingen, Tübingen, Germany; Section Computational Sensomotorics, Hertie Institute for Clinical Brain Research, Tübingen, Germany; Centre for Integrative Neuroscience (CIN), Tübingen, Germany; Department of Neurodegenerative Diseases, Hertie-Institute for Clinical Brain Research and Center for Neurology, University of Tübingen, Tübingen, Germany; Molecular Medicine, IRCCS Fondazione Stella Maris, Pisa, Italy; Koç University, Translational Medicine Research Center, KUTTAM-NDAL, Istanbul, Turkey; Groupe de recherche interdisciplinaire sur les maladies neuromusculaires (GRIMN), Centre intégré universitaire de santé et de services sociaux du Saguenay–Lac-St-Jean, Québec, Canada; Centre de recherche du Centre intégré universitaire de santé et de services sociaux du Saguenay–Lac-St-Jean, Quebec, Canada; Faculté de médecine et des sciences de la santé, Université de Sherbrooke, Québec, Canada; Department of Neurology, Heidelberg University Hospital, Heidelberg, Germany; Department of Neurodegenerative Diseases, Hertie Institute for Clinical Brain Research, Tübingen, Germany

**Author notes:** **Corresponding author:** Prof. Dr. Matthis Synofzik, Division Translational Genomics of Neurodegenerative Diseases, Hertie-Institute for Clinical Brain, Research and Center of Neurology, University of Tübingen, Hoppe-Seyler-Str. 3, 72076 Tübingen, Germany. shared last authors (Rebecca Schüle, Matthis Synofzik).

## Abstract

**Background:** With treatment trials on the horizon, this study aimed to identify candidate digital-motor gait outcomes for Autosomal Recessive Spastic Ataxia of Charlevoix-Saguenay (ARSACS), capturable by wearable sensors with multi-center validity, and ideally also ecological validity during free walking outside laboratory settings.

**Methods:** Cross-sectional multi-center study (4 centers), with gait assessments in 36 subjects (18 ARSACS patients; 18 controls) using three body-worn sensors (Opal, APDM) in laboratory settings and free walking in public space. Sensor gait measures were analyzed for discriminative validity from controls, and for convergent (i.e. clinical and patient-relevance) validity by correlations with SPRS^mobility^ (primary outcome) and SARA, SPRS and FARS-ADL (exploratory outcomes).

**Results:** Of 30 hypothesis-based digital gait measures, 14 measures discriminated ARSACS patients from controls with large effect sizes (|Cliff’s δ| > 0.8) in laboratory settings, with strongest discrimination by measures of spatiotemporal variability Lateral Step Deviation (δ=0.98), SPcmp (δ=0.94) and Swing CV (δ=0.93). Large correlations with the SPRS^mobility^ were observed for Swing CV (Spearman’s ρ = 0.84), Speed (ρ=-0.63) and Harmonic Ratio V (ρ=-0.62). During supervised free walking in public space, 11/30 gait measures discriminated ARSACS from controls with large effect sizes. Large correlations with SPRS^mobility^ were here observed for Swing CV (ρ=0.78) and Speed (ρ=-0.69), without reductions in effect sizes compared to lab settings.

**Conclusion:** We identified a promising set of digital-motor candidate gait outcomes for ARSACS, applicable in multi-center settings, correlating with patient-relevant health aspects, and with high validity also outside lab settings, thus simulating real-life walking with higher ecological validity.

## Introduction

Treatment trials are on the horizon for many spastic ataxias(1), including Autosomal Recessive Spastic Ataxia of Charlevoix-Saguenay (ARSACS) as one of the most frequent spastic ataxias worldwide (2, 3). ARSACS is a multisystemic neurodegenerative disease characterized by progressive cerebellar ataxia, spasticity, and peripheral neuropathy, with disease onset usually from early childhood to early adult years(4). Treatment trials in slowly progressive diseases like ARSACS require sensitive outcome measures that capture change in relatively short trial-like time frames (e.g. 1-2 years). While clinician-reported outcomes show only very limited sensitivity to change, sensor-based digital gait outcomes could be more sensitive, as recent findings from other genetic ataxias indicate(5).

Digital gait outcomes obtained through wearable sensors have shown promising results in other genetic ataxias, with several cross-sectional studies demonstrating reliable discrimination between patients and controls, and correlation with clinical measures of disease severity(6–8); and first longitudinal studies also demonstrating sensitivity to change(5, 9). In contrast, wearable sensor-based gait studies are still scarce in hereditary spastic paraplegias (HSPs)(10, 11), and no genotype-specific studies on sensor-based gait measures have been conducted in spastic ataxias, let alone in spastic ataxias with prominent multi-systemic involvement like ARSACS. Moreover, multi-center studies – as inevitably required for trials in such rare diseases – are very scarce in ataxias(7) and HSPs, demonstrating the urgent need for multi-center validation of performance and outcome validity. Finally, for measures to gain regulatory and patient acceptance as treatment outcomes, they must reflect health aspects that are relevant to patients(12), such as mobility(13–15). For sensor-based gait measures, this includes reflecting functional, patient-relevant disease-related impairments, ideally also with data acquired in patient-relevant settings, i.e. conditions resembling patients’ everyday lives such as e.g. public space.

To identify candidate digital motor outcomes for future trials in ARSACS, this study evaluated gait measures capturable by body-worn sensors in an international, trans-atlantic multi-center setting, assessing (i) discriminative validity against healthy controls; (ii) convergent validity with clinical measures of patient-relevant health aspects, general disease severity and activities of daily living; and (iii) ecological validity by also assessing discriminative and convergent validity in free-walking settings in public spaces.

## Methods

### Participants

The study cohort was part of the PROSPAX study, a prospective international longitudinal multi-center natural progression study in spastic ataxias (ClinicalTrials.gov, No: NCT04297891). Twenty-four patients with genetically confirmed ARSACS with available gait recordings and 50 healthy controls (HC) were recruited from four centers in four countries (Istanbul, Turkey; Pisa, Italy; Saguenay, Canada; Tübingen, Germany) based on the following inclusion criteria: (1) genetically confirmed and clinically manifest ARSACS; (2) ability to walk at least 10m freely without any walking aid; (3) absence of severe comorbidities (due to ARSACS or unrelated) which present a major confounder for evaluation of gait and stance such as: amputation, blindness, severe dementia, severe joint deformities or contractures, or fixed orthoses. HC had no history of any neurologic or psychiatric disease, no family history of neurodegenerative disease, and did not show any neurological signs upon clinical examination. After exclusion of invalid gait recordings (damaged data files, unreliable step detection) recordings of 18 subjects from the lab-based walking condition (LBW), and 15 subjects from the supervised free walking condition (SFW) remained suitable for analysis (supplementary figure 1). To ensure comparability between patients and controls, the 18 eligible patients with valid gait recordings were matched with an equal number of 18 HC such that between-group age difference was minimized by selection of the 18 youngest HC. The Institutional Review Board of the University of Tübingen approved the study (AZ 824/2019BO2). All subjects provided written informed consent before participation according to the Declaration of Helsinki.

### Clinical assessments

All participants underwent detailed neurological examination. Mobility-relevant SPRS items 1-6 were combined into a subscore termed SPRS^mobility^ (16), a clinical score to assess functional mobility across neurological systems. Disease severity was rated using the Scale for the Assessment and Rating of Ataxia (SARA) (17) and the Spastic Paraplegia Rating Scale (SPRS) (18). Activities of daily living were scored by the activities of daily living subscore of the Friedreich Ataxia Rating Scale (FARS-ADL) (19).

### Gait conditions, data acquisition, and calculation of gait measures

Walking was assessed in two different conditions: (i) constrained walking in controlled laboratory conditions(20) (laboratory-based walking, LBW) and (ii) largely unconstrained walking in public spaces, more closely resembling real-life walking (supervised free walking, SFW). See supplementary Methods 1 for detailed descriptions of gait conditions.

Sensor data were captured with three inertial sensors (Opal, APDM Wearable Technologies Inc., Portland, WA) attached on both feet and posterior trunk at the level of L5 with elastic Velcro bands.

For further details on acquisition and processing of sensor data in LBW and SFW, see supplementary Methods 2.

To obtain gait measures, non-parametric summary measures – namely median, normalized median absolute deviation (MADN = median absolute deviation/0.6745), and coefficient of variation (CV = MADN/median) – of gait features provided by Mobility Lab across strides were supplemented with one composite measure of spatial step variability (SPcmp, see below) and median harmonic ratio of pelvis linear acceleration in three directions (HR, see below). The composite measure SPcmp was formed from Stride Length CV and Lateral Step Deviation to capture spatial step variability in both anterior-posterior and medio-lateral directions in one measure, as described previously (6). To quantify gait smoothness, HR of pelvis linear acceleration was determined from raw accelerometer signals of the lumbar sensor, as described previously by us (6) and others (21).

### Selection and prioritization of gait measures

For analyzing discriminative validity, we considered a hypothesis-based selection of 30 gait measures out of all calculated measures based on previous studies and literature (from both the ataxia- and HSP-field) and clinical plausibility (6–8, 11, 22–24). A list of these 30 gait measures– including their definitions – is provided in supplementary table 1. For analyzing the convergent validity of gait sensor measures with clinical outcome assessments (COAs) in ARSACS, the correlation analyses of this study included one gait measure (out of the hypothesis-based set of 30 gait measures) of each key gait domain or subdomain established for gait disorders previously in the literature (for details of this prioritization process, see supplementary Methods 3). This led to identification of 5 gait measures, one for each gait domain or subdomain of interest: Speed (pace), Swing CV (temporal variability), Lateral Step Deviation (spatial variability), Pitch at Toe Off MADN (foot angle variability), and Harmonic ratio V (smoothness).

### Statistics and analysis

To assess the ability of gait measures to discriminate patients with ARSACS from HC, the effect size Cliff’s δ between both groups (25) was calculated for all 30 gait measures and both conditions LBW and SFW. Effect sizes were reported as small (δ ≥ 0.2), medium (δ ≥ 0.5) or large (δ ≥ 0.8) (26). Significance of group differences was determined by the non-parametric Wilcoxon ranksum test, with the significance level set to p < 0.05/n = 30: number of gait measures (Bonferroni-corrected for multiple comparisons). To assess convergent validity, Spearman correlations of prioritized gait measures with clinical- and patient-relevant COAs were examined. As primary outcome we used the patient-focused measure SPRS^mobility^, because it allows to capture patient-relevant functional mobility across neurological systems, with health aspects like gait speed or gait distance captured by the SPRS^mobility^ top-ranked by spastic ataxia patients as most relevant health aspects (Saute et al, in preparation). Exploratory outcomes were ataxia-(SARA) or spasticity-focused (SPRS) disease severity measures; and an activity of daily living scale (FARS-ADL). Effect sizes ρ were given with 95% Confidence intervals (determined by boot strapping using MATLAB’s bootci function with 2000 samples) and p-values, and classified as small (ρ ≥ 0.1), medium (ρ ≥ 0.3), large (ρ ≥ 0.5), or very large (ρ ≥ 0.7) (26, 27). Correlations with primary outcome SPRS^mobility^ were reported as significant when p < 0.05/n = 5: number of prioritized gait measures (Bonferroni corrected). Correlations with exploratory outcomes were deemed significant when p < 0.05. Further, spearman correlations between the two walking conditions (LBW, SFW) were calculated for each gait measure. Statistical analysis was performed using MATLAB (version R2022a).

## Results

Gait recordings of 18 ARSACS patients from the lab-based walking condition (LBW), and 15 ARSACS patients from the supervised free walking condition (SFW) were analysed and compared to recordings of 18 HC from the PROSPAX cohort (LBW and SFW). Individual participant characteristics are displayed in supplementary table 2.

### Analysis of gait measures discriminating between ARSACS patients and controls

To identify gait measures discriminating between ARSACS patients and HC (discriminative validity), we considered a hypothesis-based selection of 30 candidate gait measures based on previous studies, literature search and clinical plausibility. In LBW, 22 of the 30 measures discriminated ARSACS patients from HC with at least moderate effect sizes (|Cliff’s δ| > 0.5) with all of them showing significant group differences (p < 0.05/n=30: number gait measures, for overview see table 1). Fourteen measures discriminated even with large effect sizes (|δ| > 0.8). The largest effect sizes were observed for measures of spatiotemporal gait variability such as Lateral Step Deviation (δ=0.98), SPcmp (δ=0.94) and Swing CV (δ=0.93). Further measures with particularly high discriminative power (|δ| > 0.9) belonged to the category of foot angles (Pitch at Initial Contact), foot angle variability (Pitch at Toe Off MADN) and gait smoothness (Harmonic Ratio V).

**Table 1:**
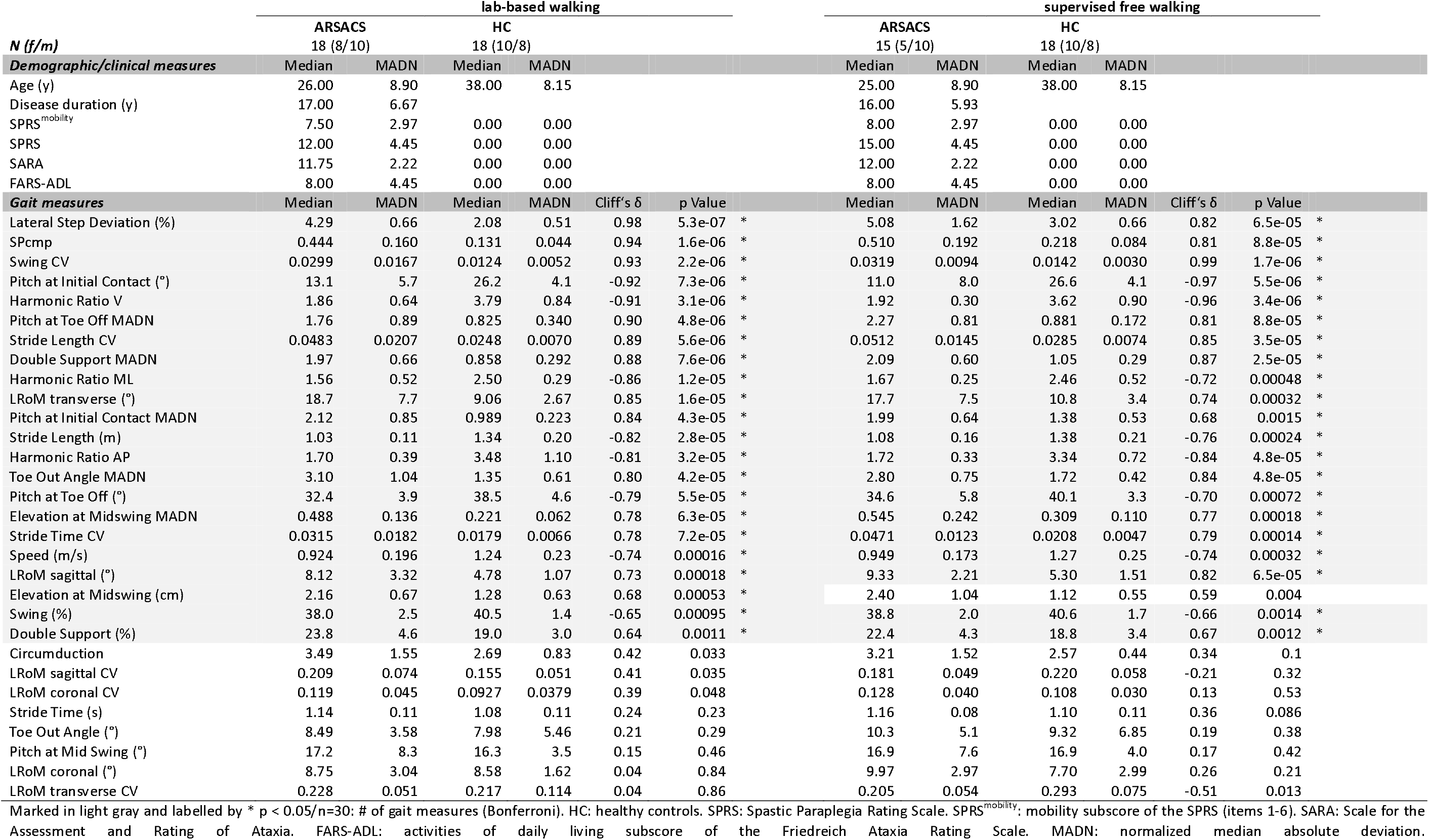
Discrimination between ARSACS patients and healthy controls in lab-based walking and supervised free walking.

| *N (f/m)* | lab-based walking |  |  |  |  |  | supervised free walking |  |  |  |  |  |
| --- | --- | --- | --- | --- | --- | --- | --- | --- | --- | --- | --- | --- |
|  | ARSACS<br>18 (8/10) |  | HC<br>18 (10/8) |  |  |  | ARSACS<br>15 (5/10) |  | HC<br>18 (10/8) |  |  |  |
| *Demographic/clinical measures* | Median | MADN | Median | MADN |  |  | Median | MADN | Median | MADN |  |  |
| Age (y) | 26.00 | 8.90 | 38.00 | 8.15 |  |  | 25.00 | 8.90 | 38.00 | 8.15 |  |  |
| Disease duration (y) | 17.00 | 6.67 |  |  |  |  | 16.00 | 5.93 |  |  |  |  |
| SPRS<sup>mobility</sup> | 7.50 | 2.97 | 0.00 | 0.00 |  |  | 8.00 | 2.97 | 0.00 | 0.00 |  |  |
| SPRS | 12.00 | 4.45 | 0.00 | 0.00 |  |  | 15.00 | 4.45 | 0.00 | 0.00 |  |  |
| SARA | 11.75 | 2.22 | 0.00 | 0.00 |  |  | 12.00 | 2.22 | 0.00 | 0.00 |  |  |
| FARS-ADL | 8.00 | 4.45 | 0.00 | 0.00 |  |  | 8.00 | 4.45 | 0.00 | 0.00 |  |  |
| *Gait measures* | Median | MADN | Median | MADN | Cliff's $\delta$ | p Value | Median | MADN | Median | MADN | Cliff's $\delta$ | p Value |
| Lateral Step Deviation (%) | 4.29 | 0.66 | 2.08 | 0.51 | 0.98 | 5.3e-07 * | 5.08 | 1.62 | 3.02 | 0.66 | 0.82 | 6.5e-05 * |
| SPcmp | 0.444 | 0.160 | 0.131 | 0.044 | 0.94 | 1.6e-06 * | 0.510 | 0.192 | 0.218 | 0.084 | 0.81 | 8.8e-05 * |
| Swing CV | 0.0299 | 0.0167 | 0.0124 | 0.0052 | 0.93 | 2.2e-06 * | 0.0319 | 0.0094 | 0.0142 | 0.0030 | 0.99 | 1.7e-06 * |
| Pitch at Initial Contact (°) | 13.1 | 5.7 | 26.2 | 4.1 | -0.92 | 7.3e-06 * | 11.0 | 8.0 | 26.6 | 4.1 | -0.97 | 5.5e-06 * |
| Harmonic Ratio V | 1.86 | 0.64 | 3.79 | 0.84 | -0.91 | 3.1e-06 * | 1.92 | 0.30 | 3.62 | 0.90 | -0.96 | 3.4e-06 * |
| Pitch at Toe Off MADN | 1.76 | 0.89 | 0.825 | 0.340 | 0.90 | 4.8e-06 * | 2.27 | 0.81 | 0.881 | 0.172 | 0.81 | 8.8e-05 * |
| Stride Length CV | 0.0483 | 0.0207 | 0.0248 | 0.0070 | 0.89 | 5.6e-06 * | 0.0512 | 0.0145 | 0.0285 | 0.0074 | 0.85 | 3.5e-05 * |
| Double Support MADN | 1.97 | 0.66 | 0.858 | 0.292 | 0.88 | 7.6e-06 * | 2.09 | 0.60 | 1.05 | 0.29 | 0.87 | 2.5e-05 * |
| Harmonic Ratio ML | 1.56 | 0.52 | 2.50 | 0.29 | -0.86 | 1.2e-05 * | 1.67 | 0.25 | 2.46 | 0.52 | -0.72 | 0.00048 * |
| LROm transverse (°) | 18.7 | 7.7 | 9.06 | 2.67 | 0.85 | 1.6e-05 * | 17.7 | 7.5 | 10.8 | 3.4 | 0.74 | 0.00032 * |
| Pitch at Initial Contact MADN | 2.12 | 0.85 | 0.989 | 0.223 | 0.84 | 4.3e-05 * | 1.99 | 0.64 | 1.38 | 0.53 | 0.68 | 0.0015 * |
| Stride Length (m) | 1.03 | 0.11 | 1.34 | 0.20 | -0.82 | 2.8e-05 * | 1.08 | 0.16 | 1.38 | 0.21 | -0.76 | 0.00024 * |
| Harmonic Ratio AP | 1.70 | 0.39 | 3.48 | 1.10 | -0.81 | 3.2e-05 * | 1.72 | 0.33 | 3.34 | 0.72 | -0.84 | 4.8e-05 * |
| Toe Out Angle MADN | 3.10 | 1.04 | 1.35 | 0.61 | 0.80 | 4.2e-05 * | 2.80 | 0.75 | 1.72 | 0.42 | 0.84 | 4.8e-05 * |
| Pitch at Toe Off (°) | 32.4 | 3.9 | 38.5 | 4.6 | -0.79 | 5.5e-05 * | 34.6 | 5.8 | 40.1 | 3.3 | -0.70 | 0.00072 * |
| Elevation at Midswing MADN | 0.488 | 0.136 | 0.221 | 0.062 | 0.78 | 6.3e-05 * | 0.545 | 0.242 | 0.309 | 0.110 | 0.77 | 0.00018 * |
| Stride Time CV | 0.0315 | 0.0182 | 0.0179 | 0.0066 | 0.78 | 7.2e-05 * | 0.0471 | 0.0123 | 0.0208 | 0.0047 | 0.79 | 0.00014 * |
| Speed (m/s) | 0.924 | 0.196 | 1.24 | 0.23 | -0.74 | 0.00016 * | 0.949 | 0.173 | 1.27 | 0.25 | -0.74 | 0.00032 * |
| LROm sagittal (°) | 8.12 | 3.32 | 4.78 | 1.07 | 0.73 | 0.00018 * | 9.33 | 2.21 | 5.30 | 1.51 | 0.82 | 6.5e-05 * |
| Elevation at Midswing (cm) | 2.16 | 0.67 | 1.28 | 0.63 | 0.68 | 0.00053 * | 2.40 | 1.04 | 1.12 | 0.55 | 0.59 | 0.004 |
| Swing (%) | 38.0 | 2.5 | 40.5 | 1.4 | -0.65 | 0.00095 * | 38.8 | 2.0 | 40.6 | 1.7 | -0.66 | 0.0014 * |
| Double Support (%) | 23.8 | 4.6 | 19.0 | 3.0 | 0.64 | 0.0011 * | 22.4 | 4.3 | 18.8 | 3.4 | 0.67 | 0.0012 * |
| Circumduction | 3.49 | 1.55 | 2.69 | 0.83 | 0.42 | 0.033 | 3.21 | 1.52 | 2.57 | 0.44 | 0.34 | 0.1 |
| LROm sagittal CV | 0.209 | 0.074 | 0.155 | 0.051 | 0.41 | 0.035 | 0.181 | 0.049 | 0.220 | 0.058 | -0.21 | 0.32 |
| LROm coronal CV | 0.119 | 0.045 | 0.0927 | 0.0379 | 0.39 | 0.048 | 0.128 | 0.040 | 0.108 | 0.030 | 0.13 | 0.53 |
| Stride Time (s) | 1.14 | 0.11 | 1.08 | 0.11 | 0.24 | 0.23 | 1.16 | 0.08 | 1.10 | 0.11 | 0.36 | 0.086 |
| Toe Out Angle (°) | 8.49 | 3.58 | 7.98 | 5.46 | 0.21 | 0.29 | 10.3 | 5.1 | 9.32 | 6.85 | 0.19 | 0.38 |
| Pitch at Mid Swing (°) | 17.2 | 8.3 | 16.3 | 3.5 | 0.15 | 0.46 | 16.9 | 7.6 | 16.9 | 4.0 | 0.17 | 0.42 |
| LROm coronal (°) | 8.75 | 3.04 | 8.58 | 1.62 | 0.04 | 0.84 | 9.97 | 2.97 | 7.70 | 2.99 | 0.26 | 0.21 |
| LROm transverse CV | 0.228 | 0.051 | 0.217 | 0.114 | 0.04 | 0.86 | 0.205 | 0.054 | 0.293 | 0.075 | -0.51 | 0.013 |

Marked in light gray and labelled by \* $p < 0.05/n=30$ : # of gait measures (Bonferroni). HC: healthy controls. SPRS: Spastic Paraplegia Rating Scale. SPRS<sup>mobility</sup>: mobility subscore of the SPRS (items 1-6). SARA: Scale for the Assessment and Rating of Ataxia. FARS-ADL: activities of daily living subscore of the Friedreich Ataxia Rating Scale. MADN: normalized median absolute deviation.

In parallel to this general exploratory analysis of 30 candidate measures, five gait measures had been prioritized a priori (see Methods), each representing a separate key gait domain in ARSACS, thus allowing to capture gait function in patients in a comprehensive, yet non-redundant manner: pace; temporal, spatial and foot angle variability; and smoothness. Four of these five measures were highly discriminative: Lateral Step Deviation, Swing CV, Harmonic Ratio V, Pitch at Toe Off MADN (|δ| ≥ 0.9), while only Speed was only moderately discriminative (δ=-0.74) (figure 1A&3C).

**Figure 1:**
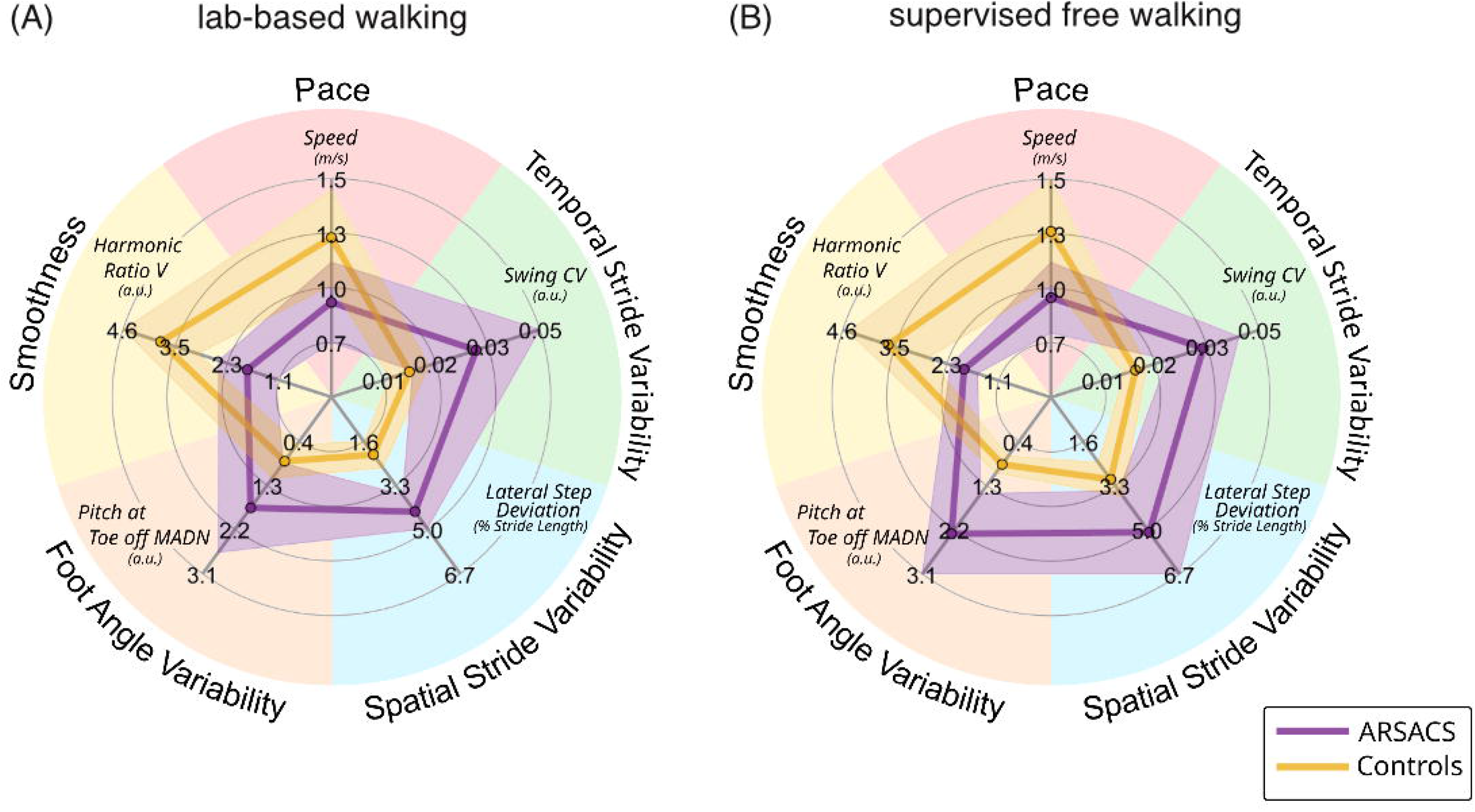
Comparison of the 5 prioritized gait measures, each representing a key gait domain in ARSACS (outer circle), between ARSACS patients and controls in (A) lab-based walking and (B) supervised free walking. Group medians ± median absolute deviations (shaded areas) for the five gait measures

### Correlation analyses of ARSACS gait measures with mobility, clinical disease severity, and activities of daily living

We assessed convergent validity of the five a priori prioritized ARSACS gait measures by examining their correlations with a set of COAs differing in their degree of functional relevance and scope (for the underlying framework see (29)). The SPRS^mobility^ had been selected as the primary outcome, given that it measures mobility at a functional level of relevance for patients, and does so across neurological systems, which is especially important in a multisystemic disease like ARSACS. Among the 5 prioritized ARSACS gait measures, the SPRS^mobility^ showed correlations of large to very large effect sizes for 3/5 measures: Swing CV (ρ=0.84, p=1.6e-5), Speed (ρ=-0.63, p=0.0050) and Harmonic Ratio V (ρ=-0.62, p=0.0066) (figure 2A, B, D, F; table 2A).

**Figure 2:**
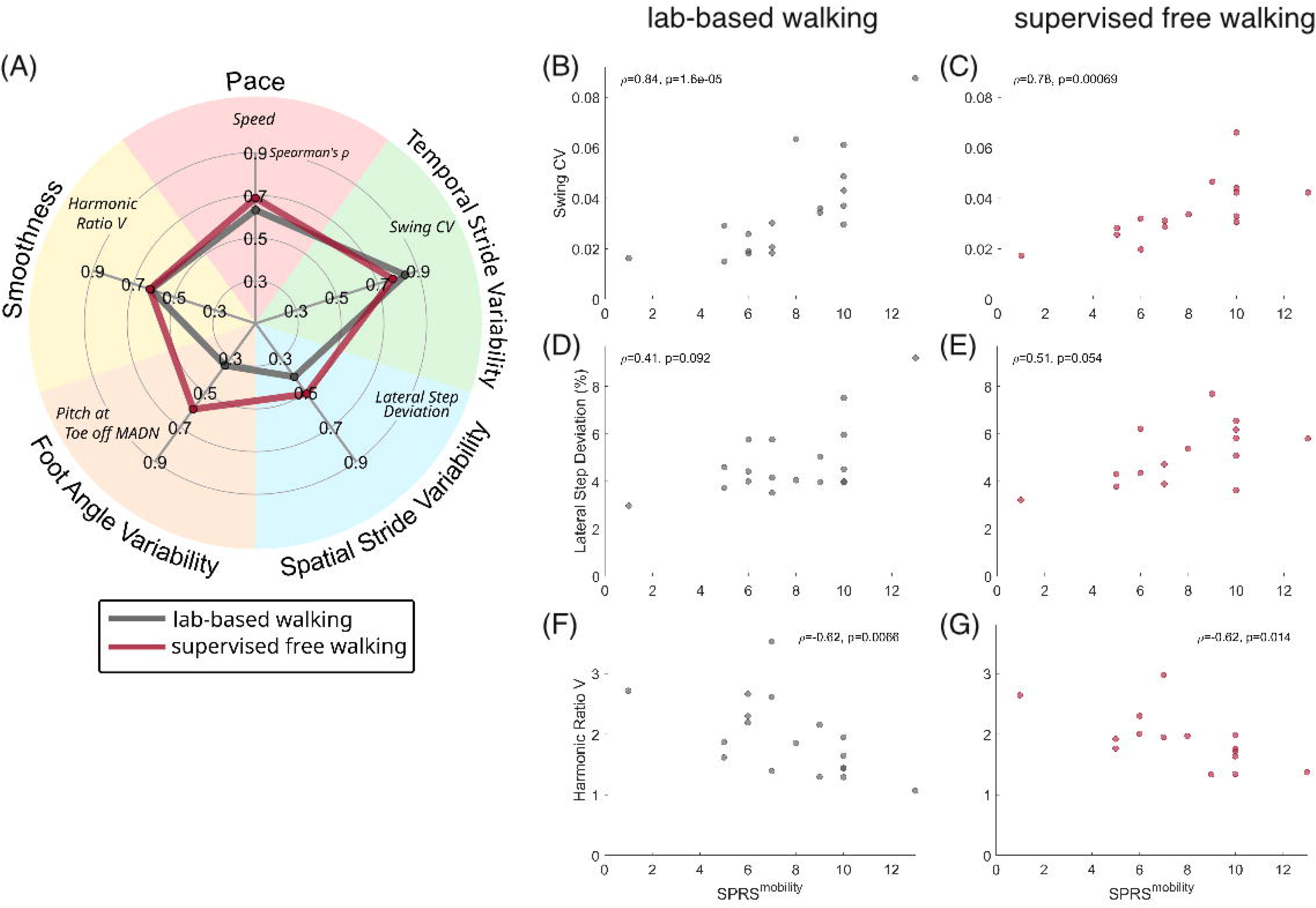
(A) Degree of correlations of the 5 prioritized gait measures, each representing a key gait domain in ARSACS (outer circle), with SPRS^mobility^ in lab-based walking (grey) versus supervised free walking (red) in ARSACS patients. (B-G). Scatter plots of three top measures versus SPRS^mobility^ in lab-based walking (left) and supervised free walking (right): Swing CV (B+C), Lateral Step Deviation (D+E), Harmonic Ratio V (F+G) .

**Table 2:** Correlation of gait measures with clinical measures in lab-based walking (A) and supervised free walking (B)

|  | **SPRS<sup>mobility</sup>** | **SPRS** | **SARA** | **FARS-ADL** |
| --- | --- | --- | --- | --- |
| **Speed (m/s)** | **-0.63 [-0.87, -0.16] \*\*** | -0.55 [-0.87, 0.12] * | -0.46 [-0.81, 0.09] | -0.03 [-0.56, 0.47] |
| **Lateral Step Deviation (%)** | 0.41 [-0.14, 0.80] | 0.37 [-0.19, 0.76] | 0.27 [-0.33, 0.72] | -0.16 [-0.64, 0.46] |
| **Swing CV** | **0.84 [0.63, 0.95] \*\*** | 0.59 [0.19, 0.84] * | 0.45 [-0.00, 0.78] | 0.49 [-0.03, 0.79] * |
| **Harmonic Ratio V** | **-0.62 [-0.85, -0.20] \*\*** | -0.45 [-0.77, 0.03] | -0.46 [-0.77, 0.09] | -0.18 [-0.65, 0.36] |
| **Pitch at Toe Off MADN** | 0.34 [-0.17, 0.72] | 0.28 [-0.35, 0.64] | 0.61 [0.22, 0.81] ** | 0.10 [-0.50, 0.57] |

**(B) Supervised free walking**
|  | **SPRS<sup>mobility</sup>** | **SPRS** | **SARA** | **FARS-ADL** |
| --- | --- | --- | --- | --- |
| **Speed (m/s)** | **-0.69 [-0.89, -0.21] \*\*** | -0.56 [-0.87, 0.01] * | 0.00 [-0.60, 0.59] | 0.08 [-0.55, 0.60] |
| **Lateral Step Deviation (%)** | 0.51 [-0.13, 0.87] | 0.41 [-0.40, 0.83] | 0.24 [-0.39, 0.74] | -0.04 [-0.64, 0.61] |
| **Swing CV** | **0.78 [0.35, 0.93] \*\*** | 0.75 [0.14, 0.94] ** | 0.30 [-0.37, 0.78] | 0.21 [-0.51, 0.72] |
| **Harmonic Ratio V** | -0.62 [-0.85, -0.19] * | -0.48 [-0.82, 0.06] | -0.28 [-0.73, 0.34] | -0.11 [-0.62, 0.46] |
| **Pitch at Toe Off MADN** | 0.60 [0.03, 0.87] * | 0.60 [-0.08, 0.92] * | 0.42 [-0.14, 0.82] | 0.09 [-0.53, 0.64] |

Spearman's $\rho$ [95% CI]. \*\* $p < 0.01$ (Bonferroni-corrected for 5 comparisons), \* $p < 0.05$
SPRS: Spastic Paraplegia Rating Scale. SPRS<sup>mobility</sup>: mobility subscore of the SPRS (items 1-6). SARA: Scale for the Assessment and Rating of Ataxia. FARS-ADL: activities of daily living subscore of the Friedreich Ataxia Rating Scale.
(A) lab-based walking
(B) supervised free walking

As exploratory outcomes we used both more symptoms-oriented clinician-reported disease severity measures focussing primarily on ataxia (SARA) or pyramidal features (SPRS); and a more general activity of daily living scale going beyond just mobility (FARS-ADL). Of the 5 prioritized ARSACS gait measures, the top measure Swing CV also correlated with activities of daily living (FARS-ADL) (ρ=0.49, p=0.040), adding further support – in addition to its correlation with the SPRS^mobility^ – that Swing CV might capture patient-relevant health-aspects. In addition to Swing CV, also the other top measure Speed correlated with the SPRS (Swing CV: ρ=0.59, p=0.010; Speed: ρ=-0.55, p=0.018), indicating an association with more general disease severity with a focus on pyramidal features. Of the 5 prioritized measures, only the foot angle variability measure Pitch at Toe Off MADN correlated with the SARA (ρ=0.61, p=0.0075) but not with the SPRS or SPRS^mobility^, suggesting a potential specificity of this measure for ataxia rather than pyramidal features.

### Analysis of gait measures discriminating between ARSACS patients and controls in free walking in public space

To evaluate gait measures under conditions with higher ecological validity – namely free walking in public space –, they were next analyzed for their discriminative power and correlation with COAs in SFW. At least large correlations between the LBW and SFW conditions were observed for 28/30 gait measures (very large: 22, large: 6, medium: 1, small: 1)(supplementary table 3). In SFW, 23/30 hypothesis-based candidate measures discriminated patients from HC with at least moderate effect sizes with 21 of them showing significant group differences. 11 measures discriminated even with large effect sizes (table 1). Significant group differences (p < 0.05/n=30: number gait measures) were observed for all measures with at least moderate effect sizes except Elevation at Mid Swing and LRoM transverse CV. The highest discriminative effect size was observed for measures of temporal variability (Swing CV, δ=0.99), foot angle (Pitch at Initial Contact, δ=-0.97) and gait smoothness (Harmonic Ratio V, δ=-0.96). Seven of the 11 measures with large effect sizes captured gait variability (temporal, spatial or foot angle). As in LBW, 4/5 a priori prioritized measures were highly discriminative Swing CV, Harmonic Ratio V, Lateral Step Deviation, Pitch at Toe Off MADN (|δ| > 0.8), while again only Speed demonstrated an only moderate effect size (δ=-0.74) (figures 1B, 3C).

**Figure 3:**
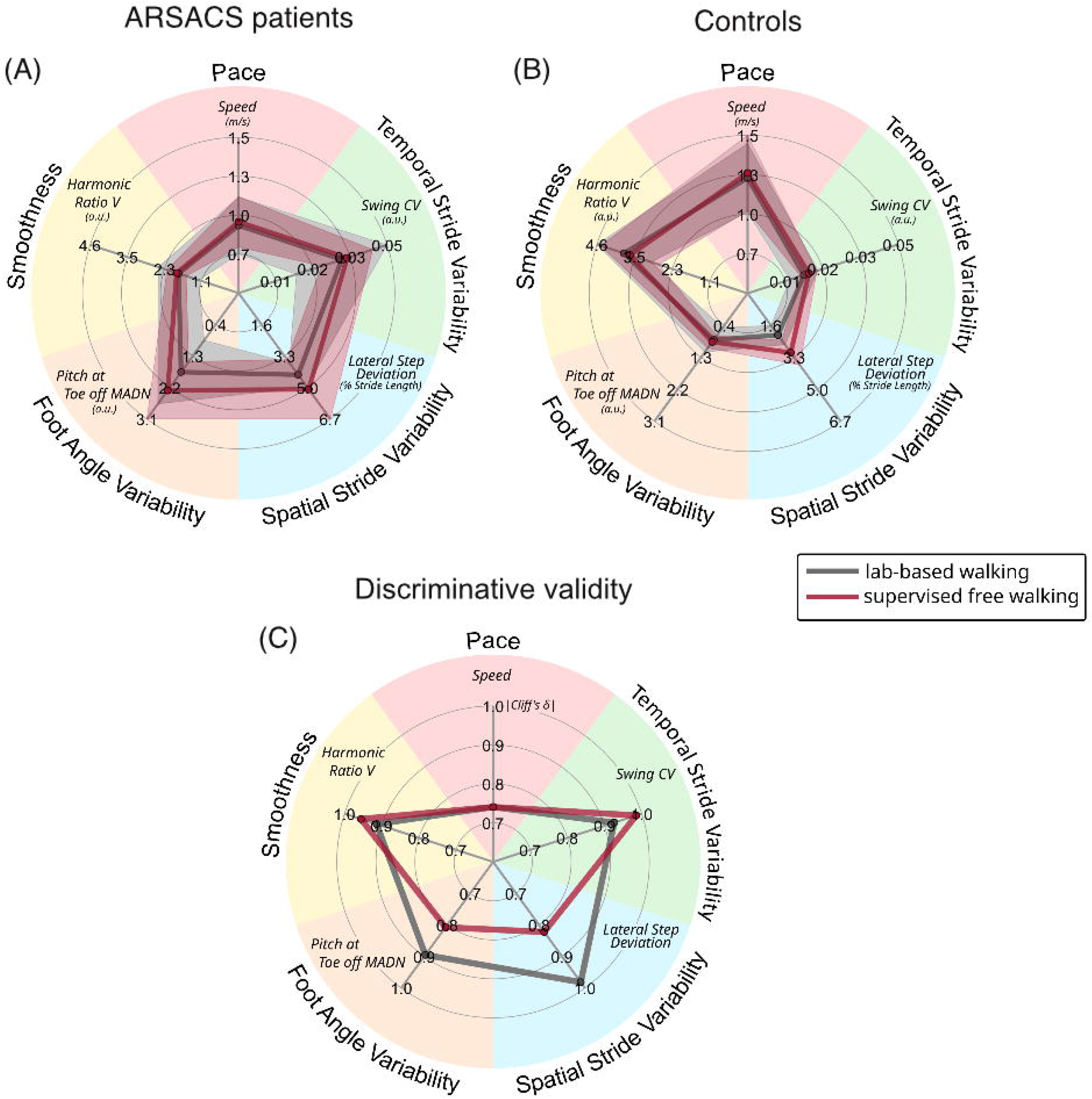
Comparison of the absolute levels of the five prioritized gait measures in (A) ARSACS patients and (B) controls between lab-based vs supervised free walking, and of (C) their discriminative effect sizes of ARSACS patients from controls in lab-based vs. supervised free walking (A, B). (A, B): group medians ± median absolute deviations (shaded areas) for ARSACS patients (A) and controls (B). (C): discriminative effect sizes Cliff’s δ of ARSACS patients versus controls.

However, compared to LBW, relevant reductions in effect size were observed in SFW for Lateral Step Deviation (SFW: 0.82; LBW: 0.98) and Pitch at Toe Off MADN (SFW: 0.81; LBW: 0.90), as depicted in figure 3C. This finding was driven primarily by two factors: relative to LBW, (i) Lateral Step Deviation in SFW increased stronger in controls than in patients; and (ii) inter-individual variance increased substantially in both Lateral Step Deviation and Pitch at Toe Off MADN in SFW, in particular in patients (figures 3A&B).

### Correlation analyses of ARSACS gait measures in free walking with clinical variables

Among the 5 prioritized ARSACS gait measures, the primary outcome SPRS^mobility^ showed correlations with large to very large effect sizes for the gait measures Swing CV (ρ=0.78, p=0.00069) and Speed (ρ=0.69, p=0.0046) also in SFW (figures 2A, C,E,G; table 2B). Compared to LBW, the effect sizes were only slightly smaller for Swing CV and even larger for Speed in SFW (figure 2A), suggesting that the effect sizes of these top gait measures observed in LBW are largely preserved even in more real-life settings with higher degrees of freedom and noise. The correlation of the SPRS^mobility^ with Harmonic Ratio V reached the same effect size in SFW as in LBW, but did formally not reach statistical significance here (ρ=-0.62, p=0.0137).

In the analysis of the exploratory outcomes, the two top measures Swing CV (ρ=0.75, p=0.0013) and Speed (ρ=-0.56, p=0.029) also correlated with the SPRS – like in LBW –, thus further underlining the association of these two measures with more general disease severity with a focus on pyramidal features. Pitch at Toe Off MADN correlated with the SPRS (ρ=0.60, p=0.017), but not with the SARA in SFW (in contrast so LBW). No significant correlations of the 5 prioritized ARSACS gait measures in SFW were observed with the SARA or the FARS-ADL.

## Discussion

This study presents the first multi-center study of digital gait outcomes in ARSACS, aiming to identify candidate digital gait outcomes for ARSACS applicable in multi-center settings and reflective of patient-relevant health aspects. With a clear focus on trial-readiness and ecological relevance, it focussed on outcomes capturable by wearable sensors demonstrating their discriminative, convergent, and ecological validity – also during free-walking in the public space.

### Gait measures discriminate patients with ARSACS from controls with large effect sizes

We identified 14 measures discriminating ARSACS patients from HC with large effect sizes (|δ| > 0.8), with the top three measures – Lateral Step Deviation, SPcmp and Swing CV – all capturing aspects of spatiotemporal stride variability. This finding corroborates and extends previous studies, demonstrating that measures of spatiotemporal stride variability are increased in ataxia, with effect sizes similar to those observed in this study (6, 7, 30); and – with, however only one study ever reported so far – also in HSP (11). Here, we demonstrate the discriminative validity of these measures the first time for ARSACS, a particular multisystemic disease combining not only ataxia and HSP features, but also severe neuropathy.

### ARSACS gait measures correlate with patient-relevant health aspects with large effect size

Specific gait measures related to temporal stride variability (Swing CV), pace (Speed), and gait smoothness (Harmonic Ratio V) revealed correlations of large effect sizes with the SPRS^mobility^, indicating convergent validity of these gait measures with a key patient-relevant COA. As opposed to classical clinician-reported scales focussing mainly on clinical symptoms, less on functions of patient relevance (like the SARA), or on general disease severity including a broad mixture of symptoms and functions (like the SPRS), the SPRS^mobility^ genuinely focusses on patient-relevant functioning. It does so by (i) capturing health aspects like gait speed or gait distance top-ranked by spastic ataxia patients as most relevant health aspects (Saute et al, in preparation); (ii) focussing on mobility-related functioning, as is key for showing convergent validity with sensor gait measures – which should inherently correlate more with mobility-related functions, rather than general disease functions (also including e.g upper limb, speech or non-motor functions); and (iii) by capturing disease-related mobility impairment across neurological systems – which is especially important in a multisystemic disease like ARSACS. Taken together, the high correlation of Swing CV, Speed, and Harmonic Ratio V with the SPRS^mobility^ indicates that these gait measures might indeed be reflective of disease-related impairment of gait and mobility, a highly patient-relevant health aspect, as shown by various patients’ voice surveys (13–15). For achieving regulatory and patient acceptance, it is mandatory for digital-motor outcomes to demonstrate that they are closely reflective of such patient-relevant health aspects, as highlighted by the recent FDA guidance (12).

Correlations in the exploratory analysis of the 5 prioritized ARSACS gait measures with the COAs SPRS, SARA and FARS-ADL exhibited smaller effect sizes, with only few reaching statistical significance. This was in accordance with our expectations, as these COAs focus mainly on clinical symptoms, less on functions of patient relevance (SARA); or on general disease severity including a broad mixture of symptoms and functions (SPRS); or on broader concepts of activities of daily living (FARS-ADL). Additionally, all of them go substantially beyond mobility, as was to be reflected by gait measures. Nevertheless, Swing CV – the measure exhibiting the largest correlation with the SPRS^mobility^ – also correlated with the FARS-ADL and SPRS. This adds further support that Swing CV might capture patient-relevant health aspects – in addition to its correlation with the SPRS^mobility^ – and even more broadly in the sense of the wide ADL spectrum captured by the FARS-ADL. It might also reflect general disease, at least as captured by a clinician-reported disease severity scale focussing on pyramidal symptoms and functioning.

### Gait measures discriminate ARSACS from controls and correlate with patient-relevant health aspects with high effect size – also in free walking in public space

Also in settings of free walking in public space (SFW), 11 gait measures with high discriminative effect size were identified, with the highest effect size again observed for measures capturing aspects of gait variability (temporal: Swing CV; spatial: Stride Length CV; foot angle: Pitch at Initial Contact) and gait smoothness (Harmonic Ratio V). This finding is remarkable as this setting – while ecologically more relevant, as closer to patients’ real life – is inherently characterized by higher degrees of freedom, noise and confounders, compared to the quiet, non-public, highly standardized lab setting (LBW). This finding is even more remarkable considering that, in addition, the walking routes in free walking naturally differed between the multiple participating centers. Indeed, a small reduction of discriminative power was observed for many measures, likely attributable to an increase of inter-individual variation in gait under free walking settings among both patients and controls that had already been observed in a previous study in other ataxia conditions (6). Yet, the same four of the five a priori prioritized ARSACS gait measures (Swing CV, Harmonic Ratio V, Lateral Step Deviation, Pitch at Toe Off MADN, all |δ| > 0.8), were highly discriminative in SFW as in LBW. Taken together, these findings validate not only the applicability of free walking test protocols in multi-center settings; but also the discriminative validity of the top-ranked ARSACS gait measures, observed in lab-based settings, in such – ecologically more relevant – free walking settings.

Moreover, similar to their performance in lab-based settings, the top gait measures related to pace (Speed), temporal variability (Swing CV), and smoothness (Harmonic Ratio V, though with p = 0.014, not statistically significant after Bonferroni-correction) also showed correlations of large to very large effect size with the SPRS^mobility^ – with no relevant decreases in effect size compared to lab settings. In fact, effect sizes for Speed were even larger in SFW than in LBW. These findings not only validate the convergent validity of the top-ranked ARSACS gait measures observed in lab-based settings, also for such free walking settings – despite the higher degrees of freedom and noise. They also demonstrate a high degree of ecological validity of these outcome measures, as they combine in fact two features of patient-relevance: these outcomes both reflect patient-relevant aspects of health (disease-related mobility), and are acquired in ecologically relevant contexts (walking in public space). This combined convergent and ecological validity of our top-ranked gait measures will be key for achieving regulatory and patient acceptance, as highlighted by the FDA guidance on patient-focussed drug development (12).

### Limitations of the study

This study has several limitations. While this study presents the first multi-center – and largest - study of any digital motor outcome in ARSACS, our findings need to be confirmed in larger ARSACS disease cohorts of still ambulatory patients. Furthermore, for some of the participants, no valid gait recordings could be obtained due to technical difficulties (unreliable step detection, corrupted data files), eligibility errors and failure in recording gait. Future studies need even more thorough eligibility and quality control training across all participating sites than already applied in the current study. Moreover, given its cross-sectional design, our study could not evaluate sensitivity to longitudinal change.

## Conclusions

This study identified a promising set of digital motor candidate gait outcomes for ARSACS, applicable in multi-center settings, that exhibited high discriminative and convergent validity in both lab-based and free walking settings, in each setting highly correlated to patient-relevant health aspects. If their sensitivity to change is validated longitudinally, these digital gait measures could serve as outcome measures in upcoming treatment trials for ARSACS and potentially also other diseases from the >200 spastic ataxias.

## Acknowledgement

This work was supported by the DFG under the frame of EJP-RD network PROSPAX (No 441409627; to M.S., R.S, F.S., C.G., A.N.B) and by the Clinician Scientist program “PRECISE.net” funded by the Else Kröner-Fresenius-Stiftung (to L.B., A.T., and M.S.). A.T. received funding from University of Tübingen, medical faculty, for the Clinician Scientist Program Grant #439–0–0. F.M.S. is supported in part by the Italian Ministry of Health (the EJP-RD network PROSPAX; Ricerca Finalizzata RF-2019-12370417; Ricerca Corrente 2023, RC 5x1000). We are grateful to Tanja Heger for monitoring the datasets of the PROSPAX registry.

## Author roles

1. Research project: A. Conception, B. Organization, C. Execution;
2. Statistical Analysis: A. Design, B. Execution, C. Review and Critique;
3. Manuscript Preparation: A. Writing of the first draft, B. Review and Critique;

L.B.: 2A, 2B, 3A

W.I.: 2A, 2C, 3C

C.K.: 1C, 3B

A.T.: 1C, 3B

S.R.: 1C, 3B

F.M.S.: 1C, 3B

A.N.B.: 1C, 3B

C.G.: 1C, 3B

R.S.: 1A, 1B, 2A, 2C, 3A, 3B

M.S.: 1A, 1B, 2A, 2C, 3A, 3B

## Additional study group contributors of the PROSPAX consortium

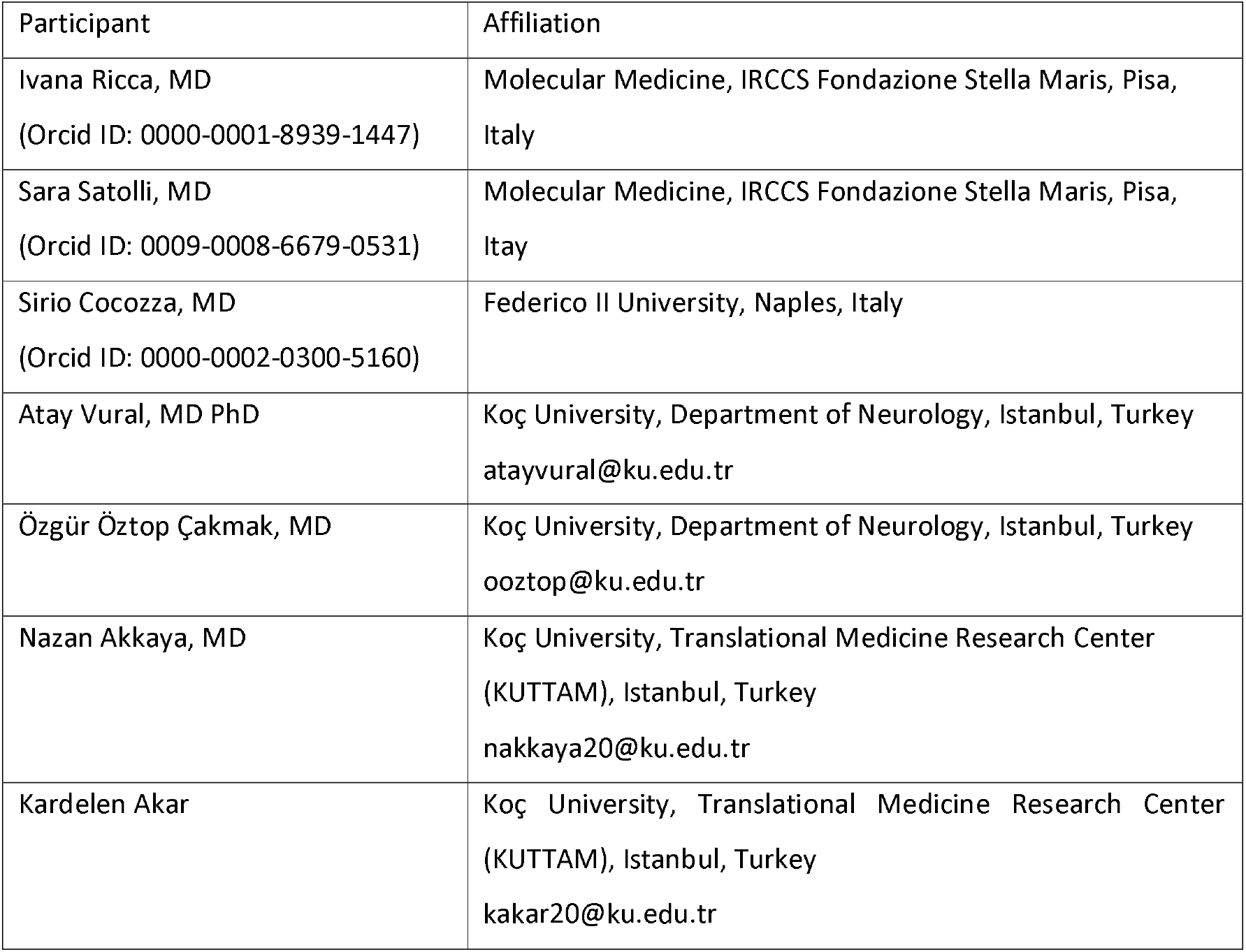

## Data availability statement

Data will be made available upon reasonable request and as patient consent allows.

## Figure Captions

**Supplementary figure 1:**
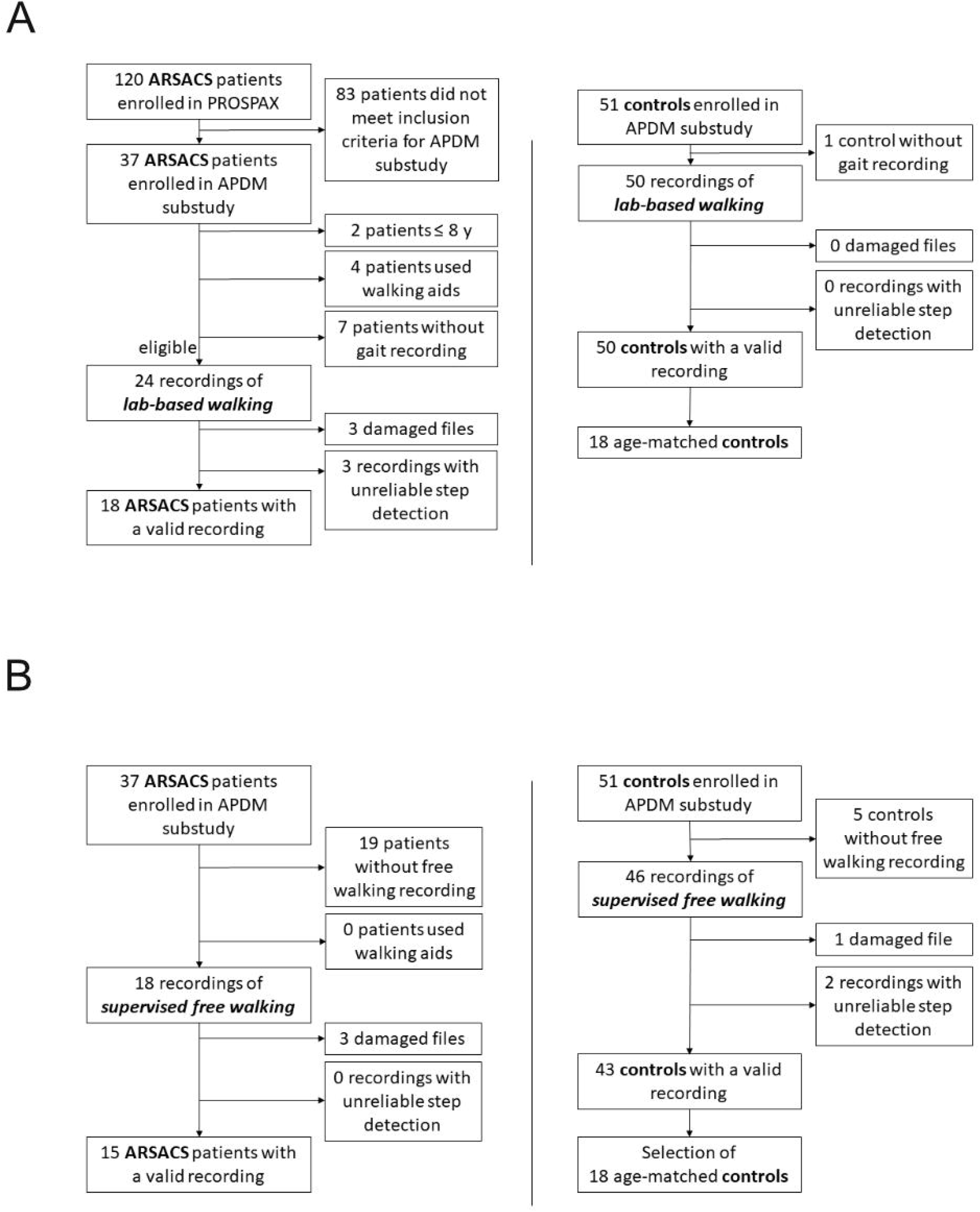
Inclusion and exclusion of gait recordings from ARSACS patients (left) and healthy controls (right) for the lab-based walking (A) and supervised free walking (B) conditions.

