## supplementary_methods for "Digital gait outcomes for ARSACS: discriminative, convergent and ecological validity in a multi-center study (PROSPAX)"

### Supplementary methods 1: Details on gait conditions

Walking movements were recorded in two different conditions:

(1) Laboratory-based walking (LBW condition): The walking distance was constrained by a specified distance of 10 meters in a specific quiet non-public indoor floor and supervised by a study assessor watching the walking performance. Participants were instructed to walk back and forth the walking distance at a self-selected speed, in accordance with Ataxia Global Initiative consensus recommendations for quantitative gait outcomes(Ilg et al., 2023). They were asked to halt after 1 minute of walking (signalled by the study assessor), and recordings were then terminated.

(2) Supervised free walking (SFW condition): Largely unconstrained walking in public spaces in an institutional (hospital) compound (all indoor: three study sites, indoor and outdoor: one study site), where participants were free to choose walking speed as they were lead along a predefined route. This walking route did include regular pedestrian encounters and bypassing, turning around corners, walking through doors and – where necessary – stairs, thus fully identical to (and part of) usual real-life walking in public spaces. Participants were supervised by a study assessor watching the participant's walking performance.

### Supplementary methods 2: Technical details on acquisition and processing of sensor data

Three Opal inertial sensors (APDM, Inc., Portland, WA) were attached on both feet and posterior trunk at the level of L5 with elastic Velcro bands. Inertial sensor data were collected and wirelessly streamed to a laptop for automatic generation of gait and balance metrics by Mobility Lab software (APDM, Inc.). For the supervised free walking condition (SFW), data were logged on board of each Opal sensor and downloaded after the session.

Step events and spatiotemporal gait features for each stride were extracted from the inertial measurement unit sensors using APDM's *Mobility Lab* software (Version 2) (Mancini et al., 2011), which has been shown to deliver good to excellent accuracy and repeatability (Morris et al., 2019; Washabaugh et al., 2017). Reliability of step detection was confirmed for each recording by inspection of a combined display of detected step events and raw accelerometer signals. In LBW, only recordings with a minimum number of 20 detected strides were included in the analysis. In SFW, only strides from walking bouts of at least 5 consecutive strides were analysed.

### Supplementary methods 3: Details on prioritization of gait measures

For analyzing the clinical validity of gait sensor measures in ARSACS, the correlation analyses of this study sought to include gait measures of each key gait domain established for gait disorders previously in the literature (Carcreff et al., 2020; Lord et al., 2013; Schniepp et al., 2020), here focussing on those gait function domains of interest for characterizing spastic-ataxic gait in neurodegenerative disease and in which corresponding measures are well capturable by APDM sensors, namely: pace, variability, and smoothness. Variability was considered in three different subdomains – temporal, spatial and foot angle (Laßmann et al., 2022; Shah et al., 2021; Velazquez-Perez et al., 2021) – given its particular interest in ataxia but also spasticity diseases, and with previous findings suggesting that these subdomains capture independent aspects of gait (Lord et al., 2013; Thingstad et al., 2015).

From the hypothesis-based set of 30 gait measures, for each of these five gait domains or subdomains (pace; temporal, spatial and foot angle variability; smoothness), we selected one gait measure per domain for correlation with clinical outcome assessments (COA) by applying the following selection criteria: avoid duplication of measures between domains; avoid redundancy of measures within the same domain; avoid measures which reflect the same gait function aspect, as indicated by too-close proximity (distance < 0.4 in both gait conditions) in the hierarchical cluster analysis (similarity: Pearson’s correlation, agglomeration: farthest neighbor) in both gait conditions. For the domain pace the measure Speed was selected because it is by far the widest accepted measure of pace in the field of neurodegenerative and neuromuscular diseases, including even gaining European Medicines Agency qualification as endpoint in trials for Duchenne muscular dystrophy (Committee for Medicinal Products for Human Use, 2023; Servais et al., 2022). For the domains variability (temporal, spatial, and foot angle) and smoothness those measures from the respective (sub-)domains were selected, that exhibited the largest effect sizes in the discrimination from healthy controls in lab-based walking.
