## supplementary_tables for "Digital gait outcomes for ARSACS: discriminative, convergent and ecological validity in a multi-center study (PROSPAX)"

**Supplementary table 1: Gait measures analyzed and their definitions**

| **Gait measures** | **Definition** | **Measure included in analysis** | | |
| --- | --- | --- | --- | --- |
|  |  | **Median** | **MADN** | **CV** |
| Stride Time (s) | The duration of a full gait cycle, measured from the left foot’s initial contact to the next initial contact of the left foot |  |  | **×** |
| Stride Length (m) | The forward distance travelled by a foot during a gait cycle | **×** |  | **×** |
| Speed (m/s) | The forward speed of the subject, measured as the forward distance travelled during gait cycle divided by gait cycle duration | **×** |  |  |
| Lateral Step Deviation  (% of Stride Length) | In a series of 3 consecutive foot placements of the same foot, the perpendicular deviations of the middle foot placement from the line connecting the ﬁrst and third, normalized by stride length | **×** |  |  |
| Pitch at Toe Off (°) | The angle of the foot as it leaves the ﬂoor at push-off. The pitch of the foot when ﬂat is zero. | **×** | **×** |  |
| Pitch at Midswing (°) | The angle of the foot at midswing. | **×** |  |  |
| Pitch at Initial Contact (°) | The angle of the foot at the point of initial contact. The pitch of the foot when ﬂat is zero and positive when the heel contacts ﬁrst. | **×** | **×** |  |
| Toe Out Angle (°) | The lateral angle of the foot during the stance phase, relative to the forward motion of the gait cycle. Positive angle is outward rotation. | **×** | **×** |  |
| Elevation at Midswing (cm) | The height of the foot sensor measured at midswing, relative to its start position while standing | **×** | **×** |  |
| Circumduction  (% of stride length) | The amount that the foot travels perpendicular to forward movement while swinging forward during an individual stride, normalized by stride length | **×** |  |  |
| Double Support  (% of gait cycle) | The percentage of the gait cycle in which both feet are on the ground | **×** | **×** |  |
| Swing (% of gait cycle) | The percentage of the gait cycle in which the foot is not on the ground | **×** |  | **×** |
| Lumbar Range of Motion, coronal (°) | The angular range of the lumbar spine in the coronal plane (roll) | **×** |  | **×** |
| Lumbar Range of Motion, sagittal (°) | The angular range of the lumbar spine in the sagittal plane (pitch) | **×** |  | **×** |
| Lumbar Range of Motion, transverse (°) | The angular range of the lumbar spine in the transverse plane (yaw) | **×** |  | **×** |
| Harmonic Ratio,  vertical (a.u.) | Harmonic ratio of pelvis linear acceleration in vertical direction, computed as described in methods | **×** |  |  |
| Harmonic Ratio,  antero-posterior (a.u.) | Harmonic ratio of pelvis linear acceleration in antero-posterior direction, computed as described in methods | **×** |  |  |
| Harmonic Ratio,  medio-lateral (a.u.) | Harmonic ratio of pelvis linear acceleration in medio-lateral direction, computed as described in methods | **×** |  |  |
| SPcmp (a.u.) | Spatial variability composite measure, computed from stride length CV and lateral step deviation as describe in methods section and elsewhere (Ilg et al., 2020) | | | |

Definitions of *Mobility Lab* gait measures from https://www.apdm.com/wp-content/uploads/2015/05/02-Mobility-Lab-Whitepaper.pdf

Ilg, W., Seemann, J., Giese, M., Traschutz, A., Schols, L., Timmann, D., & Synofzik, M. (2020). Real-life gait assessment in degenerative cerebellar ataxia: Toward ecologically valid biomarkers. *Neurology*, *95*(9), e1199-e1210. <https://doi.org/10.1212/WNL.0000000000010176>

**Supplementary table 2A: Individual participant characteristics – ARSACS patients**

| **N** | **Sex** | **Age** | **Disease Duration (y)** | **SPRS^mobility^** | **SPRS** | **SARA** | **FARS-ADL** | **No. strides (LBW)** | **No. strides (SFW)** |
| --- | --- | --- | --- | --- | --- | --- | --- | --- | --- |
| 1 | f | 41-50 | 13 | 10 | 18 | 13 | 3 | 28 | 61 |
| 2 | m | 11-20 | 12 | 8 | 12 | 11.5 | 14.5 | 25 | 392 |
| 3 | f | 21-30 | 28 | 10 | 15 | 21 | 15 | 27 | 202 |
| 4 | m | 21-30 | 20 | 5 | 10 | 15 | 14 | 32 | 105 |
| 5 | m | 41-50 | 12 | 6 | 10 | 9 | 6 | 32 | 160 |
| 6 | m | 11-20 | 8 | 1 | 6 | 4 | 1 | 29 | 316 |
| 7 | f | 31-40 | 37.8 | 7 | 12 | 8 | 9.5 | 25 | - |
| 8 | f | 21-30 | 16 | 10 | 15 | 9.5 | 11 | 30 | 178 |
| 9 | m | 21-30 | 23 | 6 | 11 | 12 | 4 | 30 | 228 |
| 10 | f | 51-60 | 13 | 13 | 18 | 14 | 8 | 22 | 43 |
| 11 | f | 11-20 | 14 | 7 | 15 | 12.5 | 8 | 26 | 126 |
| 12 | m | 31-40 | 29 | 7 | 10 | 13 | 7.5 | 24 | 96 |
| 13 | f | 11-20 | 15 | 6 | 11 | 10 | 5 | 35 | - |
| 14 | m | 21-30 | 25 | 10 | 18 | 11 | 10.5 | 39 | 251 |
| 15 | m | 31-40 | 20 | 9 | 17 | 10 | 7 | 24 | 53 |
| 16 | f | 21-30 | 25.6 | 9 | 9 | 13 | 13 | 35 | - |
| 17 | m | 11-20 | 16 | 5 | 9 | 12.5 | 6 | 29 | 188 |
| 18 | m | 21-30 | 18 | 10 | 19 | 10.5 | 16 | 29 | 160 |

**Supplementary table 2B: Individual participant characteristics – Healthy controls**

| **N** | **Sex** | **Age** |  | **SPRS^mobility^** | **SPRS** | **SARA** | **FARS-ADL** | **No. strides (LBW)** | **No. strides (SFW)** |
| --- | --- | --- | --- | --- | --- | --- | --- | --- | --- |
| 1 | m | 41-50 |  | 2 | 2 | 0 | 0 | 23 | 146 |
| 2 | m | 41-50 |  | 0 | 0 | 1 | 1 | 35 | 177 |
| 3 | f | 41-50 |  | 4 | 4 | 0 | 0 | 27 | 115 |
| 4 | m | 31-40 |  | 2 | 2 | 0 | 0 | 27 | 141 |
| 5 | m | 31-40 |  | 0 | 0 | 0.5 | 0 | 23 | 253 |
| 6 | m | 31-40 |  | 0 | 0 | 0.5 | 0 | 20 | 260 |
| 7 | m | 21-30 |  | 0 | 0 | 0 | 0 | 31 | 289 |
| 8 | f | 41-50 |  | 0 | 0 | 1 | 0 | 22 | 222 |
| 9 | f | 31-40 |  | 0 | 0 | 0 | 0 | 31 | 392 |
| 10 | f | 31-40 |  | 0 | 0 | 0 | 0 | 26 | 242 |
| 11 | f | 31-40 |  | 0 | 0 | 0 | 0 | 24 | 349 |
| 12 | f | 31-40 |  | 0 | 0 | 0 | 0 | 26 | 402 |
| 13 | m | 21-30 |  | 0 | 0 | 0 | 0 | 23 | 316 |
| 14 | f | 41-50 |  | 0 | 0 | 0.5 | 0 | 26 | 242 |
| 15 | f | 41-50 |  | 2 | 2 | 1 | 0 | 25 | 362 |
| 16 | m | 41-50 |  | 0 | 0 | 0.5 | 0 | 28 | 334 |
| 17 | f | 41-50 |  | 1 | 5 | 0 | 1 | 25 | 342 |
| 18 | f | 21-30 |  | 0 | 0 | 0 | 0 | 27 | 331 |

**Supplementary table 3: Correlation between lab-based walking (LBW) and supervised free walking (SFW) conditions**

| **Gait measure** | **Spearman’s ρ [95% CI]** | |
| --- | --- | --- |
| Circumduction | 0.96 | [0.88, 1.00] |
| Pitch at Initial Contact (°) | 0.91 | [0.53, 1.00] |
| Pitch at Mid Swing (°) | 0.86 | [0.59, 0.99] |
| Elevation at Midswing (cm) | 0.86 | [0.44, 0.99] |
| Harmonic Ratio V | 0.86 | [0.54, 0.97] |
| Pitch at Toe Off (°) | 0.85 | [0.33, 0.99] |
| LRoM coronal (°) | 0.84 | [0.56, 0.96] |
| Stride Time CV | 0.83 | [0.62, 0.97] |
| LRoM transverse (°) | 0.83 | [0.39, 0.97] |
| Toe Out Angle (°) | 0.83 | [0.54, 0.96] |
| Harmonic Ratio AP | 0.83 | [0.39, 0.96] |
| Lateral Step Deviation (%) | 0.82 | [0.61, 0.95] |
| Speed (m/s) | 0.81 | [0.50, 0.95] |
| LRoM coronal CV | 0.81 | [0.33, 0.97] |
| Stride Length (m) | 0.80 | [0.54, 0.93] |
| Toe Out Angle MADN | 0.80 | [0.41, 0.95] |
| Swing CV | 0.79 | [0.61, 0.95] |
| Swing (%) | 0.79 | [0.50, 0.95] |
| Harmonic Ratio ML | 0.79 | [0.45, 0.94] |
| Double Support (%) | 0.77 | [0.54, 0.94] |
| SPcmp | 0.73 | [0.37, 0.92] |
| LRoM sagittal CV | 0.73 | [0.22, 0.91] |
| Stride Time (s) | 0.69 | [0.09, 0.91] |
| Double Support MADN | 0.64 | [0.21, 0.89] |
| Pitch at Toe Off MADN | 0.61 | [0.20, 0.86] |
| LRoM sagittal (°) | 0.60 | [-0.24, 0.91] |
| Elevation at Midswing MADN | 0.57 | [0.06, 0.86] |
| Pitch at Initial Contact MADN | 0.50 | [-0.20, 0.91] |
| LRoM transverse CV | 0.30 | [-0.40, 0.72] |
| Stride Length CV | 0.16 | [-0.46, 0.59] |
